# Role of dietary sodium restriction in chronic heart failure. Systematic review and meta-analysis

**DOI:** 10.1101/2023.03.17.23287429

**Authors:** Szymon Urban, Michał Fułek, Mikołaj Błaziak, Katarzyna Fułek, Gracjan Iwanek, Maksym Jura, Magdalena Grzesiak, Oskar Szymański, Bartłomiej Stańczykiewicz, Kuba Ptaszkowski, Robert Zymliński, Piotr Ponikowski, Jan Biegus

## Abstract

**Background:** Dietary sodium restriction remains guidelines-approved lifestyle recommendation for chronic heart failure (CHF) patients. However, its efficacy in clinical outcome improvement is dubious.

**Methods:** We performed a systematic review of the following databases: Academic Search Ultimate, ERIC, Health Source Nursing/Academic Edition, MEDLINE, Embase, Clinicaltrials.gov and Cochrane Library (trials) to find studies analysing the impact of sodium restriction in the adult CHF population. Both observational and interventional studies were included. Most considerable exclusion criteria included i.e.: sodium consumption assessment based only on natriuresis, in-hospital interventions or mixed interventions - e.g. sodium and fluid restriction in one arm only. The review was conducted following PRISMA guidelines. At least two independent reviewers screened databases and extracted the data. Meta-analysis was performed for the endpoints reported in at least 3 papers. The heterogeneity and sensitivity of the results were assessed. Analyses were conducted in Review Manager (RevMan) Version 5.4.1 The Cochrane Collaboration, 2020.

**Results:** Initially, we screened 9175 articles identified in the databases. Backward snowballing revealed 1050 additional articles. Eventually, 9 papers were evaluated in the meta-analysis. All-cause mortality, HF-related hospitalizations, and the composite of mortality and hospitalization were reported in 8, 6 and 3 articles, respectively. Sodium restriction was associated with a higher risk of composite endpoint (OR, 4.12 [95% CI, 1.23 - 13.82]) and did not significantly affect the all-cause mortality (OR, 1.38 [95% CI, 0.76 - 2.49]) or HF hospitalisation (OR, 1.63 [95% CI, 0.69 - 3.88]).

**Conclusions:** In a meta-analysis, sodium restriction in CHF patients worsened the prognosis in terms of composite of mortality and hospitalizations and did not influence all-cause mortality and HF hospitalisation rate.

## Introduction

For many years, sodium restriction has been recommended as a key dietary intervention for patients with heart failure (HF)^1^. This recommendation was based on the belief that a low-sodium diet would reduce fluid retention and decrease the risk of HF hospitalizations^2^. While, this assumption was based on the fact that in HF, water and sodium homeostasis are greatly disturbed and any interference may lead to clinical deterioration. However, recent studies have cast doubt on the effectiveness of this intervention, with some suggesting that it may even be harmful^3^.

One of the main criticisms of sodium restriction in the management of HF is that the evidence supporting its implementation is rather weak and based on early experiments that involved assessing the pathological responses of HF patients to sodium loading^4^. Thus, these experiments may not accurately reflect the effects of long-term sodium restriction. On the other hand, the burden of pharmacological and non-pharmacological recommendations in HF is significant and often challenging to maintain in the long run^5–7^.

Given these unclear or conflicting perspectives, there is a significant need to evaluate the effect of recommended low sodium diet in HF. In our meta-analysis, we aim to provide a comprehensive evaluation of the existing evidence on the benefits and risks of sodium restriction in the management of HF.

## Methods

### Search strategy

Initially, we screened the following databases: Academic Search Ultimate, ERIC, Health Source Nursing/Academic Edition, MEDLINE, Embase, Clinicaltrials.gov, and Cochrane Library (trials) for the relevant articles. No restrictions regarding the publication date were determined, screening and papers export were performed on 11.10.2022. The keywords differed slightly in different sources.

In EMBASE, Academic Search Ultimate, ERIC, Health Source Nursing/Academic Edition and Cochrane Library, keywords were as follows: ((diet*) OR (eat*) OR (ingestion) OR (feed) OR (micronutrient) OR (macronutrient) OR (intake*) OR (nutri*) OR (consump*)) AND ((heart failure) OR (ventricular dysfunction) OR (HF) OR (HFpEF) OR (HFrEF) OR (cardiomyopat*) OR (((cardia*) OR (myocardial)) AND ((failure) OR (insufficienc*)))) AND ((sodi*) OR (salt)). In the CochraneLibrary, trials section was screened for suitable papers, and Cochrane Reviews were screened for the reviews available for the backward snowballing, but non-relevant reviews were identified. Searching in Clinicaltrials.gov included the following conditions: Condition or disease: ((heart failure) OR (ventricular dysfunction) OR (HF) OR (HFpEF) OR (HFrEF) OR (cardiomyopat*) OR (((cardia*) OR (myocardial)) AND ((failure) OR (insufficienc*)))) Other terms: ((diet*) OR (eat*) OR (ingestion) OR (feed) OR (micronutrient) OR (macronutrient) OR (intake*) OR (nutri*) OR (consump*)) OR ((sodi*) OR (salt)). Only studies with the status of completed, terminated or unknown were screened. Studies which analysed children were excluded from the search engine.

All the records (n=9175) were exported into the excel file, and duplicates were removed (n=395); further, two independent reviewers (S.U., M.F.) screened the titles, abstracts and full-texts.

In the second step, backward snowballing was performed by 2 reviewers (O.S. and M.G.). Both references of included articles and papers which cited them were screened for the relevant records. Reviews and editorials were also screened^1,2,8–10^, and 1050 records were identified.

### Eligibility criteria

Inclusion criteria were defined as: studies analysing the impact of dietary sodium restriction on HF patients’ outcome, full-text, peer-reviewed articles written in English, the population of heart failure patients of age> 18 years, both interventional and observational studies, reporting of at least one of the endpoint of interest: all-cause, cardiovascular or HF-related mortality; all-cause, cardiovascular or HF-related hospitalisation, emergency department visit, HF decompensation.

Exclusion criteria were as follows: studies analysing the amount of the added salt (not the total dietary sodium restriction), case reports or review articles, studies based on the animal models, mixed interventions, e.g. studies that limited both sodium and fluids only in the intervention arm, in-hospital intervention only, and studies in which the time of the intervention was shorter than the follow-up time.

Studies which assessed the sodium consumption based only on the natriuresis were excluded, as it was shown that such a method of sodium consumption evaluation is unsatisfactory in the patients treated with loop diuretics^11.^ The review was performed following PRISMA guidelines^12^ and was registered in PROSPERO (CRD42023391133).

### Data collection and analysis

After the screening, data extraction was performed by 2 independent reviewers (K.F. and M.G.), and the discrepancies were solved by the discussion with the input of the third investigator (S.U.). Authors of the papers with missing data necessary for the quantitative analysis were contacted to obtain relevant information. Data regarding study design, inclusion and exclusion criteria, sample size, age and sex of participants, method of sodium consumption assessment, amount of sodium restriction, length of follow-up, mortality, HF hospitalisations, composite endpoint compounds and occurrence and serum creatinine levels were extracted. Studies with no events in both arms were not included in the meta-analysis^13^.

We used random effect with the Mantel-Haenszel test to analyse categorical variables. The odds ratio and 95% confidence intervals (CI) were calculated for the outcomes. Continuous variables were assessed using the inverse variance method and random effects model. Heterogeneity was evaluated using I^2^ (I^2^>50% was considered significant heterogeneity). Funnel plots for the publication bias assessment and meta-regression were not performed due to the limited number of included studies^13,14^. Review Manager version 5.4.1 (The Cochrane Collaboration, 11-13 Cavendish Square, London, W1G 0AN United Kingdom) was used for the statistical analysis.

### Subgroup and sensitivity analysis

Trial-level subgroup analysis was performed to investigate the source of the heterogeneity. We assessed the effect in the following subgroups: follow-up longer and shorter than 1 year; heart failure with reduced ejection fraction (HFrEF) population only and merged HFrEF and heart failure with preserved ejection fraction (HFpEF); and sodium restriction below 2 g per day. Sensitivity analysis, which included randomised control trials and observational studies separately, was performed for the selected outcomes.

### Risk of bias assessment

The risk of bias (ROB) in the selected studies was assessed using Cochrane-designed tools. Risk of Bias 2 and ROBIN-I were used for randomised trials and observational studies respectively^15,16^. Two independent reviewers (S.U. and O.S.) performed a quality evaluation, and the discussion resolved all the discrepancies. The study was considered low risk when the ROB was assessed as a low risk in all the domains.

### Grading the quality of evidence

The overall quality of the acquired evidence was evaluated using the Grading of Recommendations Assessment, Development, and Evaluation (GRADE) approach^17^. Each outcome was assessed separately for randomised and observational studies. GradePRO GDT (McMaster University and Evidence Prime Inc.) was used to create a Summary of Findings and Certainty of Evidence table.

## Results

The results of the search are displayed in Figure 1. The research revealed 9 articles, including 2210 participants - 1130 in the sodium-restricted (SR) and 1080 in the sodium-liberal (SL) group. Five studies were RCTs, 2 prospective cohort studies and 2 propensity score matching registry analyses. Patients were observed from 12 weeks to 3 years (mean follow-up time 15.77 months). Three studies included the HFrEF population exclusively, while 6 studies analysed the subjects regardless of the ejection fraction. In 5 studies, researchers restricted sodium consumption to 2 g per day or less, and 5 studies included only patients with accurately chosen NYHA class. Men constituted 64% of patients, and the mean age was 67 years. The characteristic of the included studies is described in Table 1.

**Table 1.**
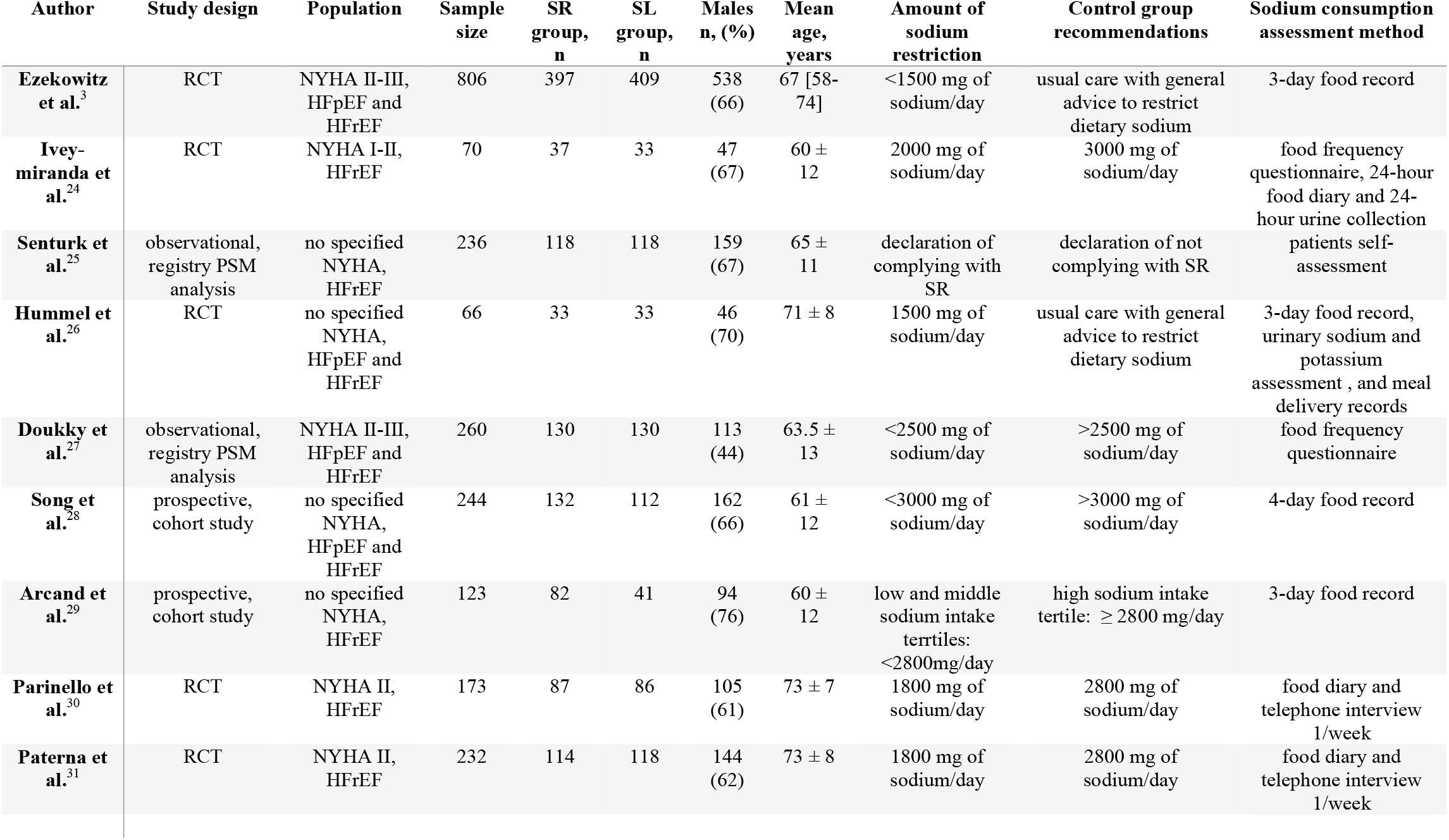
Characteristic of the included studies. Per protocol data are shown – intended values of sodium restriction and the number of enrolled patients are displayed. Age is presented as mean *±* standard deviation, except study by Ezekowitz et al. – median [interquartile range]. Abbreviations: SR – sodium restriction, SL – sodium liberal, RCT – randomised controlled trials, PSM – propensity score matching, NYHA – New York Heart Association, HFpEF – heart failure with preserved ejection fraction, HFrEF – heart failure with reduced ejection fraction.

**Figure 1.**
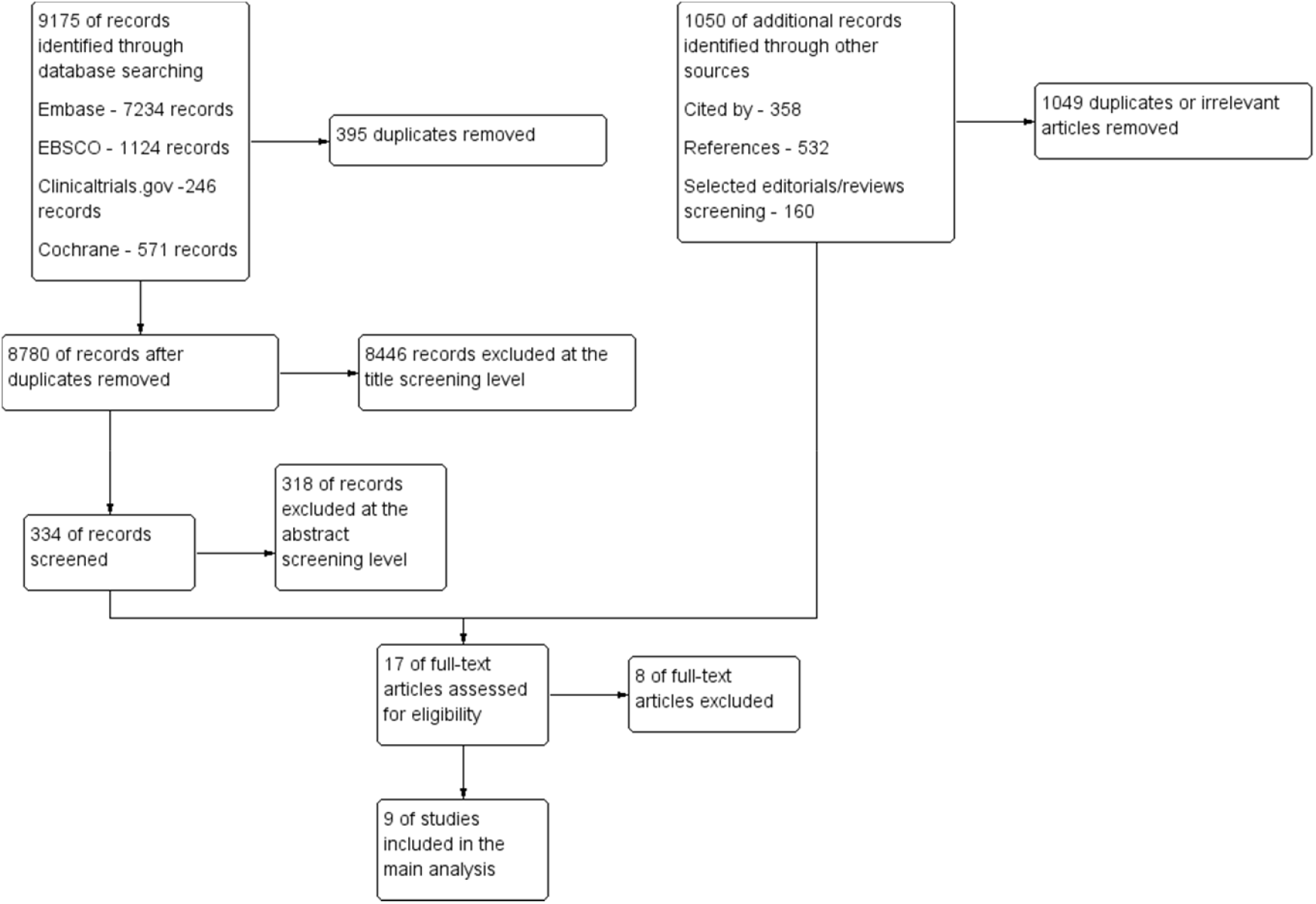
Flowchart of the systematic review process. Excel files with the screened articles and exclusion reasons at every stage are available at request.

Four parameters, i.e. all-cause mortality, HF hospitalisation, a composite of mortality and readmission and serum creatinine level, were reported in at least 3 studies; therefore, quantitative analysis of these features was performed. A summary of the findings is shown in Table 2. Forrest plots of the merged analyses are displayed in Figure 2. Specific subgroup data with the accurate number of patients with events are available in the Supplementary material (Figure S1).

**Table 2.** Summary of findings of the included studies. Mean follow-ups are presented in the observational studies. Abbreviations: SR – sodium restricted, SL – sodium liberal, NR – not reported, SD – standard deviation.

| Author | Duration of follow-up, months | Mortality n, (%) | HF hospitalization n, (%) | Composite of death and hospitalization n, (%) | Creatinine mg/dl, mean ± SD |
| --- | --- | --- | --- | --- | --- |
| Ezekowitz et al. | 12 | SR 22 (6); SL 17 (4) | NR | NR | NR |
| Ivey-Miranda et al. | 5 | NR | NR | SR 8 (21); SL 7 (21) | SR 1.16 ± 0.06; SL 1.20 SD 0.06 |
| Senturk et al. | 20 | SR 35 (10); SL 36 (31) | SR 70 (59); SL 87 (74) | NR | NR |
| Hummel et al. | 3 | SR 0 (0); SL 1 (3) | SR 7 (21); SL 13 (39) | NR | SR 1.4 ± 0.4; SL 1.3 SD 0.5 |
| Doukky et al. | 36 | SR 24 (19); SL 14 (11) | SR 42 (32); SL 26 (20) | NR | NR |
| Song et al. | 12 | SR 3 (2); SL 3 (3) | SR 14 (11); SL 8 (7) | NR | NR |
| Arcand et al. | 28 | SR 2 (2); SL 6 (15) | NR | NR | NR |
| Parinello et al. | 12 | SR 20 (23); SL 4 (5) | SR 44 (51); SL 12 (14) | SR 64 (73); SL 16 (19) | SR 2.1 ± 0.5; SL 1.45 SD 0.4 |
| Paterna et al. | 6 | SR 15 (13); SL 6 (5) | SR 30 (26); SL 9 (8) | SR 45 (39); SL 15 (13) | SR 2.1 ± 0.5; SL 1.45 SD 0.4 |

**Table 4.**
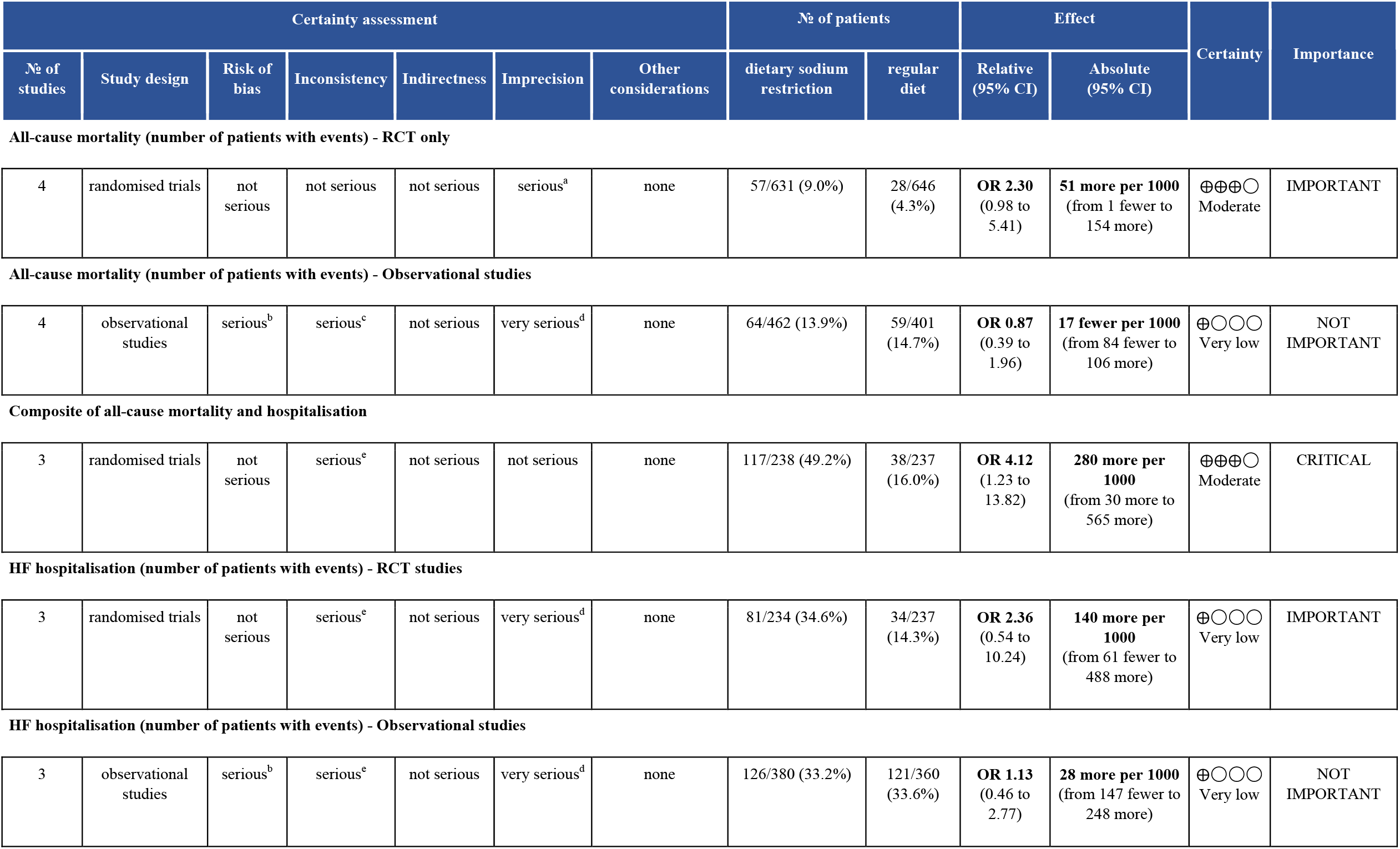
Summary of Findings and Certainty of Evidence Table. Explanations CI: confidence interval; OR: odds ratio a. Number of events does not meet the optimal information size of 400 events, 95% CI includes no effect and appreciable harm. b. Some limitations for multiple criteria, mainly serious bias in the selection of participants into the study in all studies and serious bias due to confounding in half of the studies. c. Substantial inconsistency that can be partially explained as a result of different cut off for the amount of sodium in sodium reduced diet as well as different participation of HFpEF. d. The number of events does not meet the optimal information size of 400 events, 95% CI includes no effect and appreciable benefit and harm. e. Considerable inconsistency that can be partialy explained as a result of different cut off for amount of sodium in sodium reduced diet as well as different participation of HFpEF.

| Certainty assessment |  |  |  |  |  |  | № of patients |  | Effect |  | Certainty | Importance |
| --- | --- | --- | --- | --- | --- | --- | --- | --- | --- | --- | --- | --- |
| № of studies | Study design | Risk of bias | Inconsistency | Indirectness | Imprecision | Other considerations | dietary sodium restriction | regular diet | Relative (95% CI) | Absolute (95% CI) |  |  |
| All-cause mortality (number of patients with events) - RCT only |  |  |  |  |  |  |  |  |  |  |  |  |
| 4 | randomised trials | not serious | not serious | not serious | serious<sup>a</sup> | none | 57/631 (9.0%) | 28/646 (4.3%) | **OR 2.30**<br>(0.98 to 5.41) | **51 more per 1000**<br>(from 1 fewer to 154 more) | ⊕⊕⊕○<br>Moderate | IMPORTANT |
| All-cause mortality (number of patients with events) - Observational studies |  |  |  |  |  |  |  |  |  |  |  |  |
| 4 | observational studies | serious<sup>b</sup> | serious<sup>c</sup> | not serious | very serious<sup>d</sup> | none | 64/462 (13.9%) | 59/401 (14.7%) | **OR 0.87**<br>(0.39 to 1.96) | **17 fewer per 1000**<br>(from 84 fewer to 106 more) | ⊕○○○<br>Very low | NOT IMPORTANT |
| Composite of all-cause mortality and hospitalisation |  |  |  |  |  |  |  |  |  |  |  |  |
| 3 | randomised trials | not serious | serious<sup>c</sup> | not serious | not serious | none | 117/238 (49.2%) | 38/237 (16.0%) | **OR 4.12**<br>(1.23 to 13.82) | **280 more per 1000**<br>(from 30 more to 565 more) | ⊕⊕⊕○<br>Moderate | CRITICAL |
| HF hospitalisation (number of patients with events) - RCT studies |  |  |  |  |  |  |  |  |  |  |  |  |
| 3 | randomised trials | not serious | serious<sup>c</sup> | not serious | very serious<sup>d</sup> | none | 81/234 (34.6%) | 34/237 (14.3%) | **OR 2.36**<br>(0.54 to 10.24) | **140 more per 1000**<br>(from 61 fewer to 488 more) | ⊕○○○<br>Very low | IMPORTANT |
| 3 | observational studies | serious<sup>b</sup> | serious<sup>c</sup> | not serious | very serious<sup>d</sup> | none | 126/380 (33.2%) | 121/360 (33.6%) | **OR 1.13**<br>(0.46 to 2.77) | **28 more per 1000**<br>(from 147 fewer to 248 more) | ⊕○○○<br>Very low | NOT IMPORTANT |

**Figure 2.**
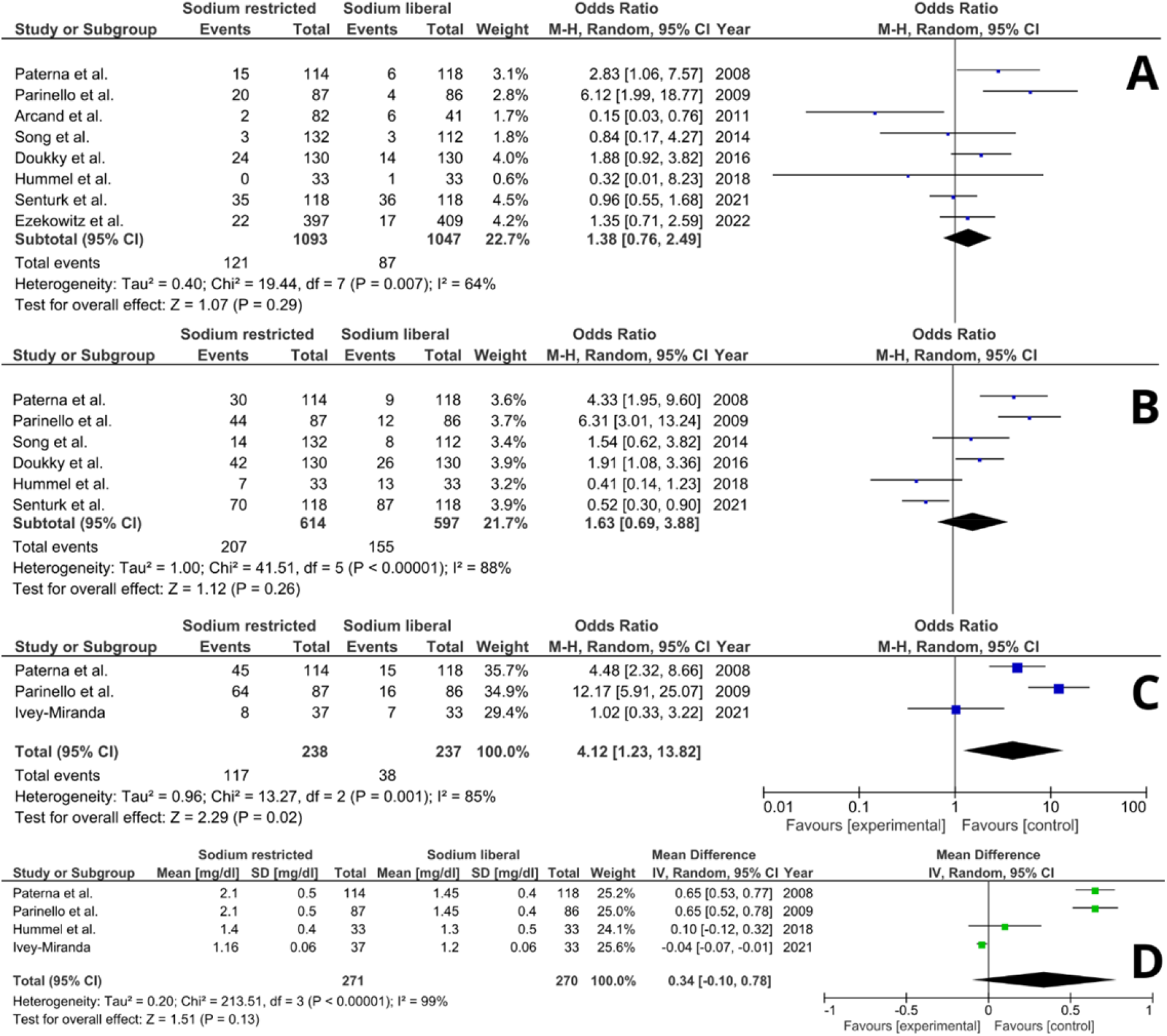
Forrest plots of the sodium restricted vs sodium liberal diet for the analysed outcomes. Forrest plots of the subgroup analyses are available in Supplement Figure S1. A-all-cause mortality; B – HF hospitalization; C – composite of mortality and readmission, D – serum creatinine level.

### All-cause mortality

All-cause mortality was reported in 8 studies with 2140 participants. The death occurred in 121 (11.07%) in the SR group and 87 (8.31%) in the SL group. Dietary sodium restriction did not significantly affect mortality in HF patients (OR, 1.38 [95% CI, 0.76 - 2.49]). It was close to significance in the RCTs analysis, reaching (OR, 2.30 [95% CI, 0.98 - 5.41], p=0.06). The effect remained neutral in subgroup and sensitivity analysis: (OR, 0.87 [95% CI, 0.39 – 1.96]) in observational studies, (OR, 1.28 [95% CI, 0.65 –2.50]) and (OR, 1.69 [95% CI, 0.28 – 10.35]) in studies with > 1-year and < 1-year follow-up respectively, (OR, 1.86 [95% CI, 0.93 – 3.75]) in studies which restricted sodium consumption to 2 g per day, (OR, 1.51 [95% CI, 0.24 – 9.67]) and (OR, 1.45 [95% CI, 0.92 – 2.28]) in studies which included HFrEF only and in HFpEF and HFrEF respectively.

### HF hospitalisations

HF-related hospitalisations occurrence was assessed in 6 studies with 1211 patients. The endpoint took place in 207 (33.71%) patients in SR and 155 (25.96%) in the SL population. Sodium restriction did not significantly affect the HF hospitalisations rate (OR, 1.63 [95% CI, 0.69 – 3.88]). The comparable results have been accomplished in the analysis of the following subgroups: RCTs (OR, 2.36 [95% CI, 0.54 – 10.24]); observational studies (OR, 1.13 [95% CI, 0.46 – 2.77]); follow up longer (OR, 1.74 [95% CI, 0.61 – 4.95]) and shorter (OR, 1.38 [95% CI, 0.14 – 13.74]) than 1 year; sodium restriction equal or below 2 g/d (OR, 2.14 [95% CI, 0.68 – 6.77]); HFrEF analysis only (OR, 2.38 [95% CI, 0.45 – 12.66]) and merged HFpEF and HFrEF (OR, 1.13 [95% CI, 0.45 – 2.83]).

### Composite endpoint

The composite endpoint of all-cause mortality and hospitalisation was analysed in 3 papers (475 participants). The composite endpoint occurred in 117 (49.16%) patients in SR and 38 (16.04%) in SL cohorts. A diet with a restricted amount of administered sodium significantly increased the risk of death or hospitalisation (OR, 4.12 [95% CI, 1.23 – 13.82]). Due to the low number of studies, no subgroup analysis regarding this outcome was performed.

### Serum creatinine level

Serum creatinine level at the end of the follow-up was reported in 4 studies (541 participants). The mean difference between the groups was estimated as 0.34 mg/dL [95% CI, -0.1 – 0.78]) suggesting that serum creatinine may be higher in the SR population. The results, however, did not reach statistical significance (p=0.13).

### Risk of bias and certainty of the evidence

The results of the ROB assessment are displayed in Figure 3. The quality of evidence assessed by GRADE criteria for the observational studies was very low. The quality of the evidence in RCTs was: moderate for all-cause mortality, very low for HF hospitalisation and moderate for a composite of mortality and hospitalisations (Table 3).

**Figure 3.**
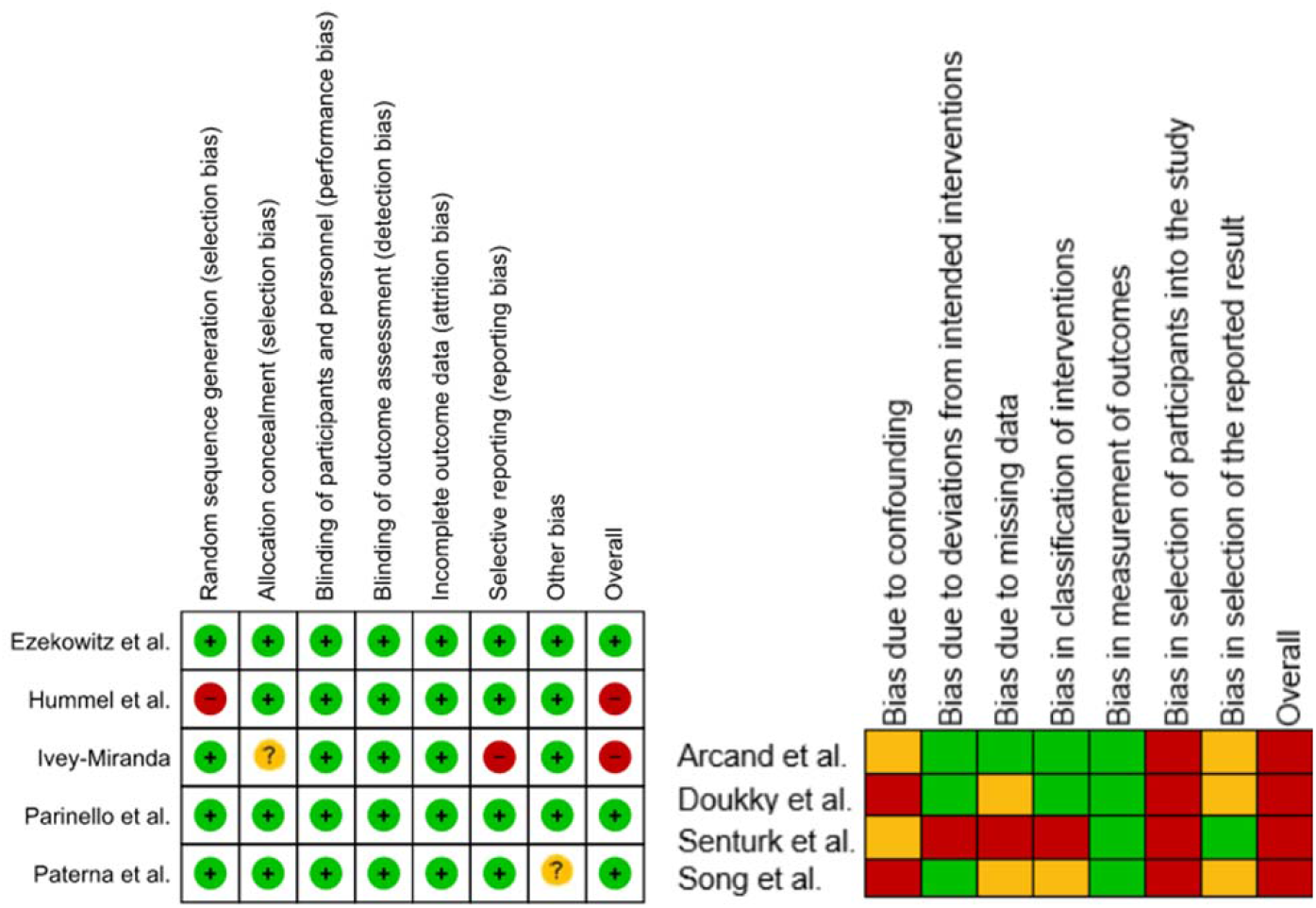
Risk of bias (ROB) in the included studies. ROB was assessed separately for the randomised (left side) and observational studies (right side). Cochrane tools, i.e. risk of bias 2 for randomised trials and ROBINS-I for the observational studies, were used. Red colour marks high ROB, green low ROB. The yellow colour describes the risk as unclear in randomised data and moderate in observational.

## Discussion

In this interventional and observational data meta-analysis, we evaluated current evidence for dietary sodium restriction in the HF population. Our data showed that the sodium restriction does not provide benefit in terms of outcome improvement. Importantly these results remained neutral regardless of the: type of study (RCT vs observational), left ventricular ejection fraction, duration of follow-up and amount of sodium restriction. Further, sodium restriction showed no benefit in any of the analysed outcomes. Its impact on all-cause mortality and HF hospitalisations was insignificant, while it was meaningful in terms of the composite of mortality and hospitalisation. Sodium restriction significantly increased the risk of the composite endpoint (OR, 4.12 [95% CI, 1.23 – 13.82], p=0.02).

Noteworthy, our analysis is the first one to show the aggregated impact of sodium restriction on serum creatinine levels, which seems to be higher in the SR group, without reaching statistical significance (+ 0.34 mg/dL [95% CI, -0.1 – 0.78], p=0.13). This finding may be partly explained by the prognostic role the increased natriuresis plays in the decongestion process^18^. Further studies are warranted to elucidate the phenomenon comprehensively.

SODIUM-HF^3^ was the most numerous RCT that analysed the effect of dietary sodium restriction in HF. Its results have considerably questioned the long-lasting, guidelines-supported paradigm. Following the trial, the authors prepared a meta-analysis of the existing evidence on that topic^8^. Our results complement the works mentioned above; nevertheless, some essential differences require clarification. Firstly, opposite the authors, we did not include studies that analysed salt and fluid restriction in one of the arms. While the fluid restriction is also being questioned as a standard of care^19,20^, its conjunction with sodium restriction may affect the results. We have also analysed only outpatient interventions. Finally, we have not included studies which based the sodium consumption assessment exclusively on natriuresis, as it was shown to be inappropriate in the patients administering loop diuretics^11^.

Omitting its possible harmful effect, the assumption that the sodium restriction in HF may be unjustified has profound clinical implications. HF patients receive a long list of recommendations, some of which are troublesome and inconvenient for the patients^5^. Presuming that the patients have the limited ability to cover all the physicians’ advice, as we take care of the patient’s compliance, we should focus on communicating and underlining indications with a reliable, evidence-based effect. In the same manner, as physicians should fight polipharmacy^21^, they should try to eliminate unreasonable recommendations that may distract patient attention from the essential ones.

Our study is not free from limitations. The scheduled analysis design, which included both interventional and observational studies, was, per se, associated with the higher ROB. Different scales are designed to assess the ROB in the observational and interventional studies^22,23^, which makes it difficult to compare the reliability of the included studies. Moreover, analysed studies were published between 2008 and 2022 – during this period, 4 European and American guidelines were published. Patients’ management, e.g. recommended pharmacotherapy, did change remarkably, which may impact the generalizability of the results. Further, the analysed studies presented considerable heterogeneity - they differed considerably in terms of design, e.g. time of follow-up, included population, amount of sodium restriction and concomitant interventions. Finally, there was an inherent problem with blinding the participants and the personnel due to the nature of the studied, nutritional intervention.

In conclusion, dietary sodium restriction seems not to prevent HF patients from death or HF - hospitalisation, with some low-quality evidence about its association with adverse effects. Given that, there is no sufficient evidence that dietary sodium restriction should be recommended for the general HF population anymore.

## Data Availability

Excel files with the screened articles and exclusion reasons at every stage are available at request. All the extracted data will be shared upon request.

